# Evaluating ChatGPT’s Responses to Chiropractic Patient Questions: A Comparative Study

**DOI:** 10.64898/2026.10.25.26366458

**Authors:** Alex V. Pham, Alexander C. Peterson, Jessica C. Tom, Aubrianna L. Jones, Clinton J. Daniels

## Abstract

**Background:** Large language models (LLMs) are used at an increased frequency by patients to answer health related questions. Prior studies have evaluated LLMs and their performance with musculoskeletal (MSK) and spine domains, but have not been used to evaluate chiropractic-relevant content.

**Objective:** To evaluate ChatGPT responses to commonly asked chiropractic patient questions for concordance with chiropractic clinical practice guidelines.

**Methods:** Thirty questions addressing low back pain, neck pain, and chronic musculoskeletal pain were developed based on current U.S. chiropractic clinical practice guidelines (CPGs). ChatGPT (GPT-5.2) generated responses in isolated sessions. Responses were assessed for readability using established indices, then five licensed chiropractors independently rated each response using a 5-point Likert scale across four domains: relevance, clarity, completeness, and accuracy. Inter-rater reliability was evaluated using Krippendorff’s alpha (ordinal).

**Results:** Readability measures demonstrated that ChatGPT responses were written at approximately a college-level reading grade. Expert ratings across all domain means (standard deviation) were predominately clustered at the upper end of rating scale (1-5): Relevance 4.72 ± 0.56; Clarity 4.76 ± 0.50; Completeness 4.23 ± 0.81; Accuracy 4.00 ± 1.02. The Krippendorff’s alpha (ordinal) ranged from -0.023-0.214 across the domains, all reflective of poor inter-rater reliability.

**Conclusion:** ChatGPT responses were broadly consistent with chiropractic CPGs, though readability levels exceeded recommended patient education standards. While the expert panel frequently assigned similar scores, minor scoring differences and restricted score distributions produced low chance-corrected reliability. These findings suggest that ChatGPT can produce guideline concordant responses but readability and interpretive variability should be considered when integrating LLM generated information into patient education.

## Introduction

The internet provides a near limitless amount of information that is easily accessible to the public. Approximately 75% of Americans report utilizing online resources for medical advice.^1^ While useful for ease of access, the quality of information remains varied and unregulated, which raises concerns for the accuracy and safety of consumed medical advice. Since the inception of artificial intelligence (AI) chat bots, and more recently conversational chatbots using large language models (LLMs) (i.e., ChatGPT), information has become increasingly easily available and these programs have demonstrated the capability to generate human-like responses and even the ability to pass professional exams.^2^ The ChatGPT LLM has been demonstrated to provide accurate and empathetic answers to patient questions.^3^ However, other studies have identified critical limitations of LLMs, including producing incorrect or irrelevant responses.^4–6^ The quality of responses are often dependent on how a prompt is phrased by a user prior to entry.^7^ The user limitations of LLMs highlight that continued progress is needed and warrants evaluation of LLM generated content against verified standards.^8,9^

Similar to social media, concerns have been raised that the rapid expansion of generative AI has the potential for LLMs may create or perpetuate misinformation as related to orthopedic conditions and chiropractic treatments.^10–13^ The United States’ low back pain clinical practice guidelines (CPG) emphasize conservative, non-pharmacological care as first-line therapies^14,15^, and chiropractic profession CPGs highlight the chiropractor’s role in patient education, encouraging lifestyle modifications, and clarifying patient concerns.^16–18^ Clinical practice guidelines are important for patient safety and it is important that public-facing resources reflect best practices.^19,20^ To date, no studies have examined ChatGPT’s responses to patient questions about chiropractic health care and whether they are consistent with current chiropractic CPGs.

Previous research in similar domains (physical therapy and spine care) suggest that ChatGPT can meet 80% compliance with musculoskeletal guidelines.^8,21^ It remains to be seen if this pattern will remain similar with chiropractic specific queries, which may present unique topics such as the safety of spinal manipulation therapy, appropriate use of diagnostic imaging, or a chiropractic provider’s clinical assessment and decision-making process.^22–28^.

Given the rise of consumer-facing LLMs, understanding their performance in a chiropractic context has scientific, clinical, and ethical implications. Therefore, the purpose of this study will be to investigate the quality of ChatGPT responses to questions related to chiropractic. The primary aim was to investigate ChatGPT responses to chiropractic-related questions for readability, relevance, clarity, completeness, and accuracy with clinical guidelines.

## Materials and Methods

### Study Design

This study builds on prior work assessing ChatGPT’s responses to patient questions regarding epidural steroid injections and musculoskeletal rehabilitation.^7,8^ This study assessed ChatGPT’s responses to chiropractic-related questions from a patient perspective. For each question, responses from ChatGPT were compared to recent CPG recommendations related to chiropractic management of neck pain, mechanical low back pain, and chronic musculoskeletal pain.^16–18^ The study protocol was reviewed by the VA Puget Sound Human Research Protection Office and determined to be exempt from comprehensive Institutional Review Board (IRB) review.

### Query Development

Thirty commonly asked chiropractic questions were identified and categorized into three categories: *1) low back pain, 2) neck pain, and 3) general musculoskeletal pain*. (Table 1) The selection of questions for ChatGPT queries were derived to align with statements from recent U.S. CPGs about low back pain, neck pain, and chronic musculoskeletal pain.^16–18^ Questions were then reviewed by all study investigators for clarity and alignment with topics addressed in current U.S. CPGs.^16–18^

**Table 1.**
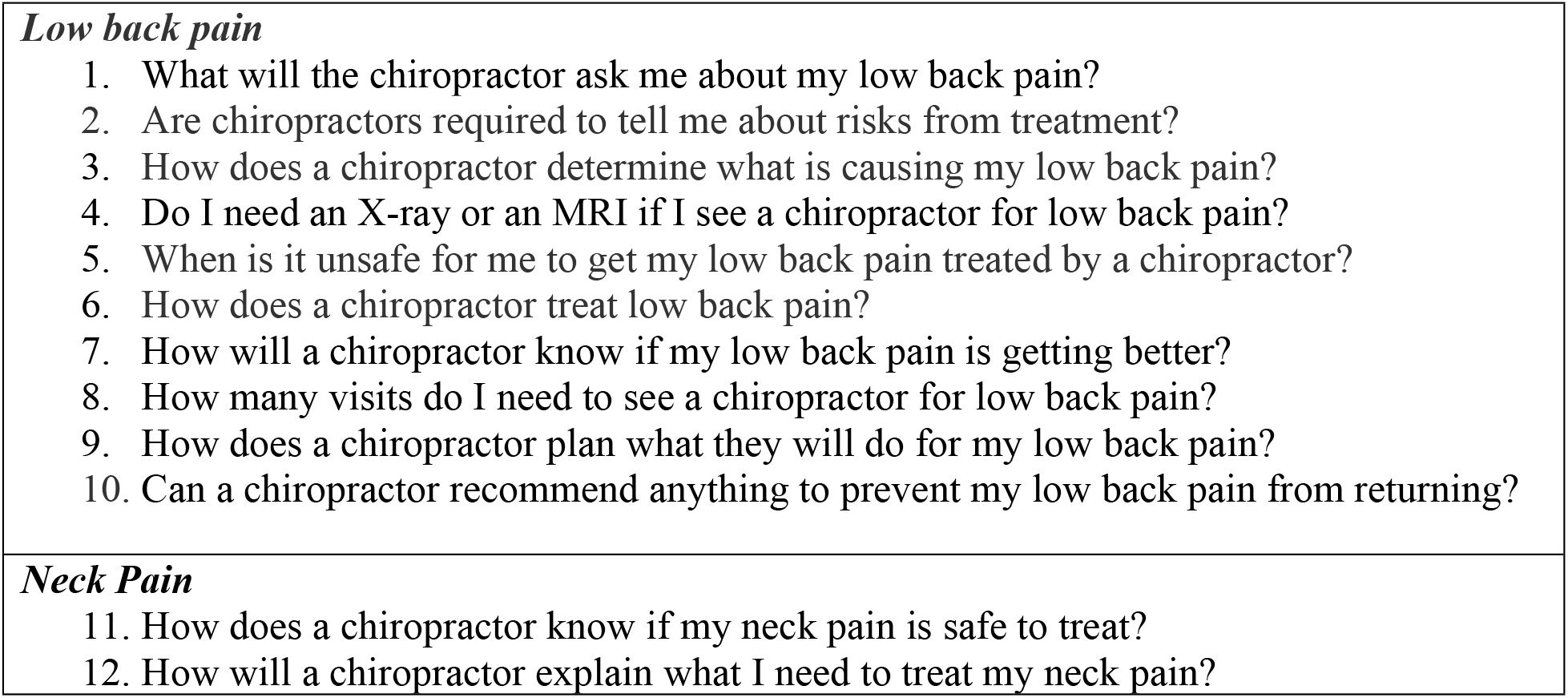
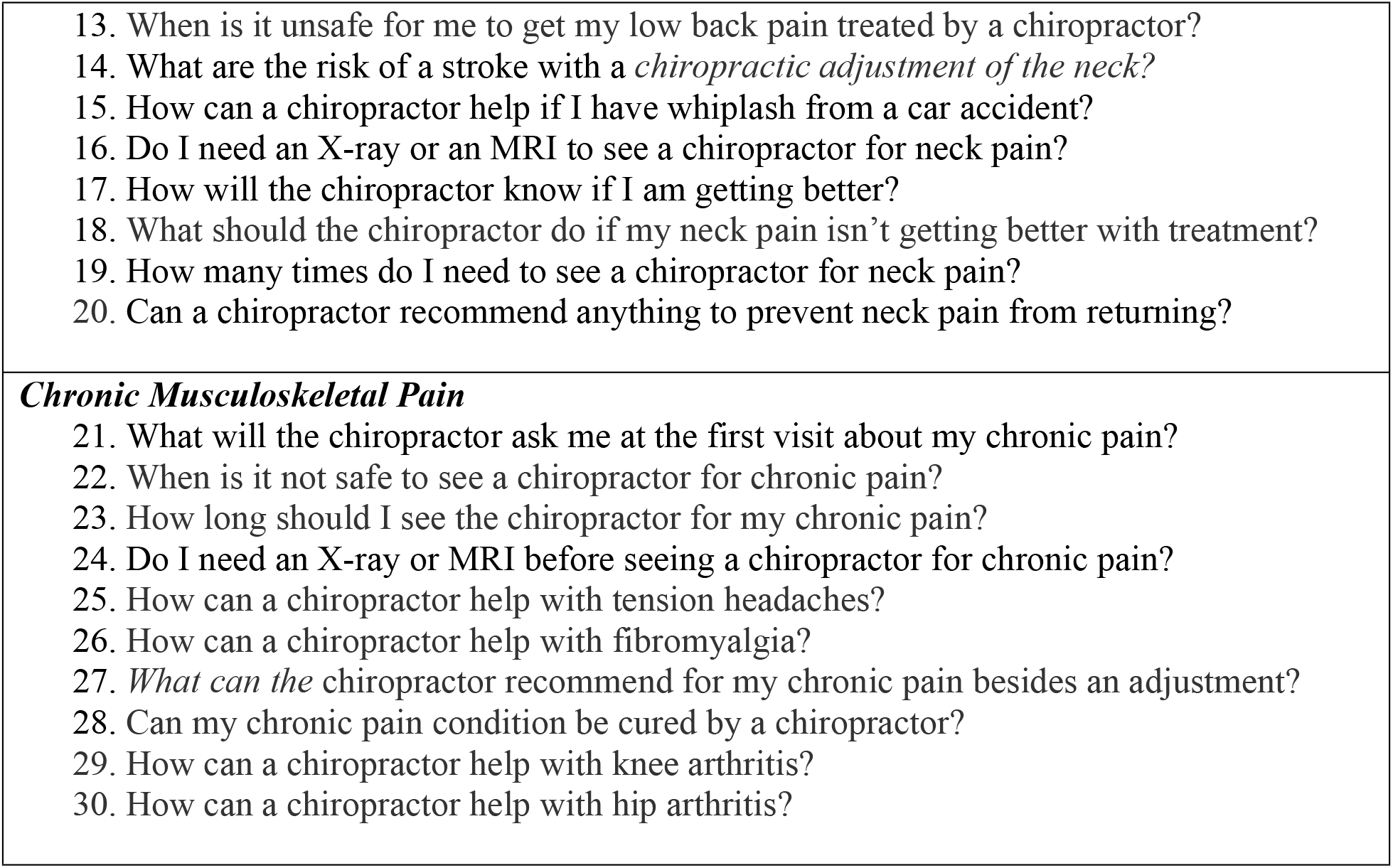
Questions used to query ChatGPT.

### ChatGPT Response Generation

The OpenAI website ChatGPT (GPT-5.2) was utilized in December 2025 without signing into an account. Each question was input in separate conversations, with the preceding conversation being deleted prior to the submission of the next question. The model was not given any prompt instruction and responses were recorded verbatim.

### Statistical Analysis

To assess *Readability*, the Repostseo Readability Score Checker online program was used to assess structural components of responses. Descriptive statistics (mean, standard deviation) were reported for word count, character count, number of sentences, words per sentence, characters per word, Flesch-Kincaid grade level, and Flesch Reading Ease score.

A panel of five subject matter experts completed a structured 5-point Likert scale (1 – strongly disagree, 2 – disagree, 3 – neither agree or disagree, 4 – agree, 5 – strongly agree) across the four domains of *Relevance, Clarity, Completeness*, and *Accuracy* (Table 2). The five-member subject matter expert panel was comprised of licensed chiropractors with clinical, educational, and research experience. Each panelist independently reviewed all thirty ChatGPT-generated responses, scoring each domain individually.

**Table 2.**
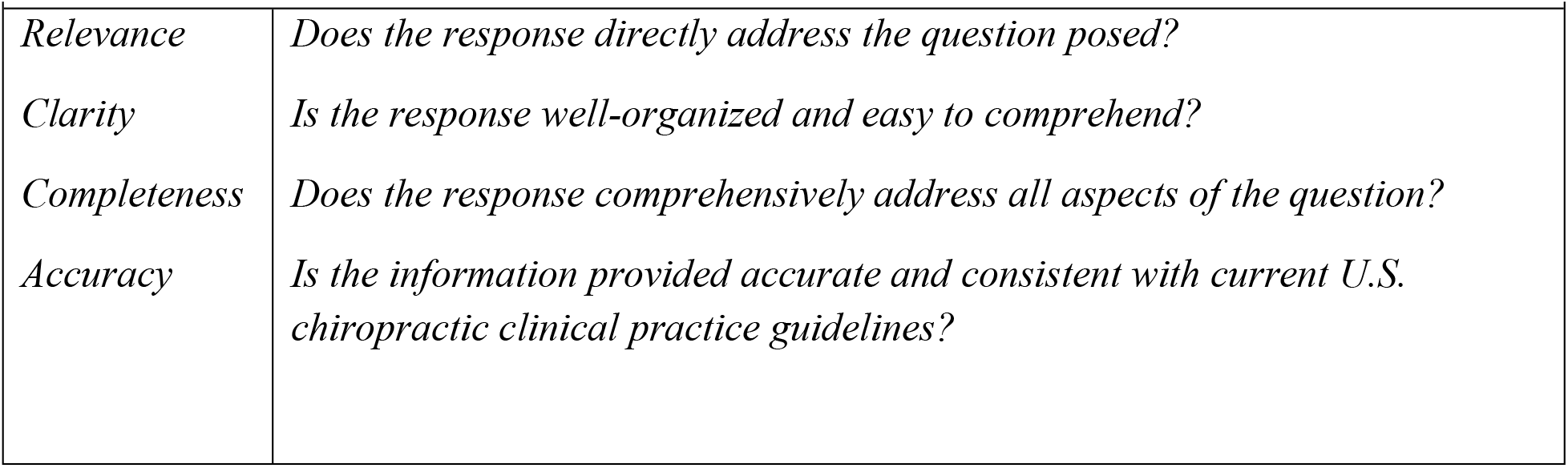
Definitions.

| *Table 2. Definitions* |  |
| --- | --- |
| *Relevance* | *Does the response directly address the question posed?* |
| *Clarity* | *Is the response well-organized and easy to comprehend?* |
| *Completeness* | *Does the response comprehensively address all aspects of the question?* |
| *Accuracy* | *Is the information provided accurate and consistent with current U.S. chiropractic clinical practice guidelines?* |

Likert scores were reported as means and standard deviations. To assess *Accuracy* the panelist compared the ChatGPT responses to verbatim statements from the CPGs. All statistical analysis will be conducted through Microsoft Excel, with the exception of Krippendorff’s Alpha (ordinal) which was calculated using the K-Alpha Calculator^29^

## Results

The ChatGPT responses and paired clinical practice guideline are available in Supplementary Files A and B. The response structure breakdown is presented in Table 3. Flesch-Kincaid Grade level and Flesch Reading Ease score indicated the written responses were consistent with an advanced level that is suitable for college graduates.

**Table 3.** Response structured components.

| *Table 3. Response structured components* |  |
| --- | --- |
| **Query Component** | **Response (mean, SD)** |
| Word Count | 279.03 (50.01) |
| Character Count (no spaces) | 1693.17 (321.49) |
| Sentence Count | 11.03 (3.00) |
| Words per sentence | 27.35 (9.11) |
| Characters per word | 6.08 (0.31) |
| Flesh-Kincaid Grade Level | 13.61 (3.61) |
| Flesch Reading Ease | 8.34 (12.22) |

Subject matter experts scored the domains of Relevance and Clarity the highest, followed by Completeness, and Accuracy was scored the lowest (Table 4). The means of individual questions are presented in Table 5, with question 3 “*How does a chiropractor determine what is causing my low back pain?”* rated the highest across all domains and response to question 23 “*How long should I see the chiropractor for my chronic pain*?” rated the lowest. Responses to low back pain questions were generally rated higher than responses for neck pain or general musculoskeletal pain. (Table )

**Table 4.** Domain Ratings Across 30 items for five raters.

| *Table 4. Domain Ratings Across 30 items for five raters* |  |  |  |  |
| --- | --- | --- | --- | --- |
| **Domain** | **Mean ± SD** | **Median (IQR)** | **% Rated 4–5** | **Range** |
| Relevance | 4.72 ± 0.56 | 5 (5-5) | 95% | 3-5 |
| Clarity | 4.76 ± 0.50 | 5 (5-5) | 97% | 3-5 |
| Completeness | 4.23 ± 0.81 | 4 (4-5) | 82% | 1-5 |
| Accuracy | 4.00 ± 1.02 | 4 (3-5) | 71% | 1-5 |

**Table 5.**
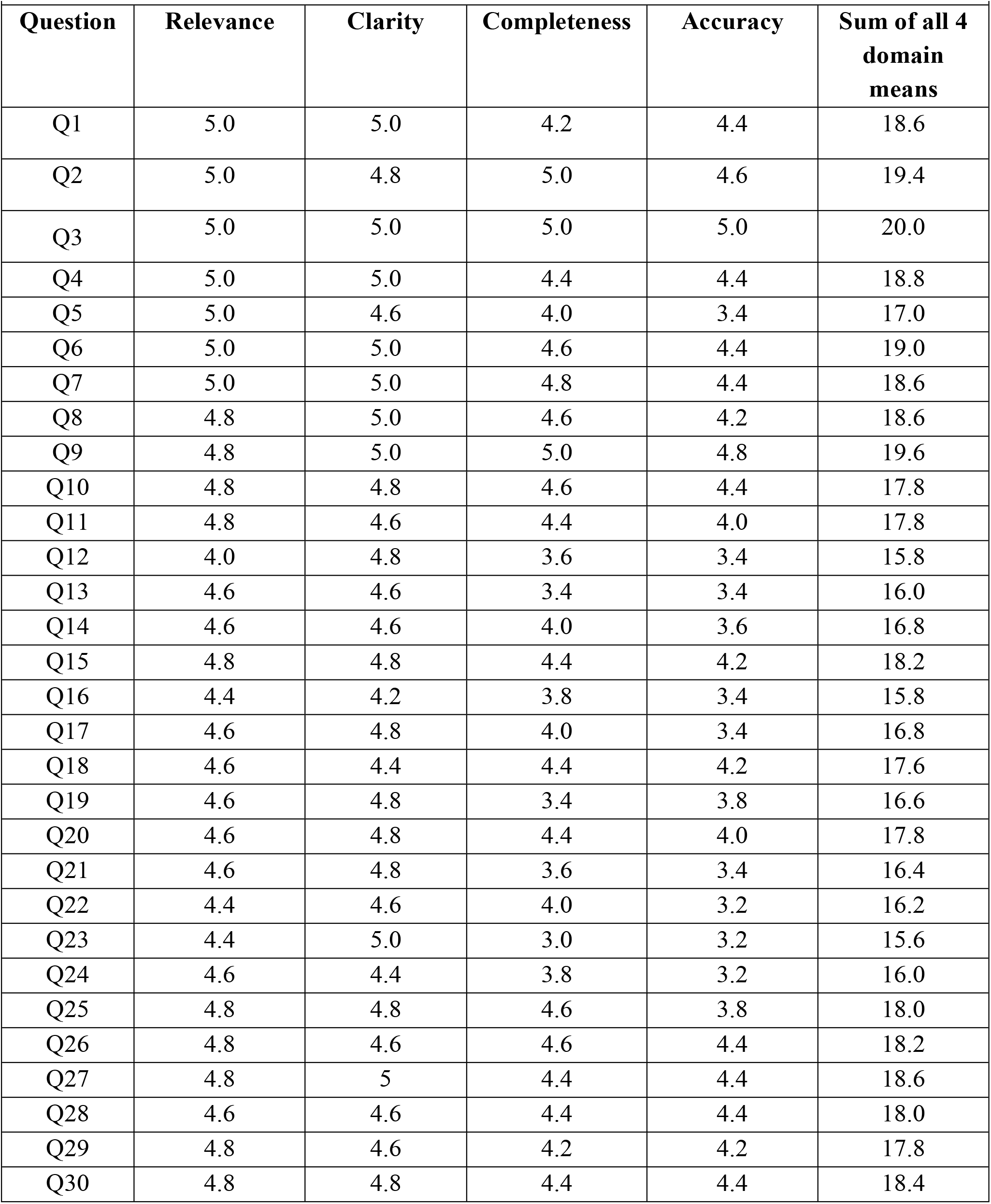
Comparison mean score of five expert review of relevance, clarity, completeness and accuracy.

**Table 5a.** Category-wise mean score of five expert reviewers.

| *Table 5. Category-wise mean score of five expert reviewers* |  |  |  |  |
| --- | --- | --- | --- | --- |
| **Category** | **Relevance** | **Clarity** | **Completeness** | **Accuracy** |
| Low back pain (Q1-10) | 4.94 | 4.92 | 4.62 | 4.40 |
| Neck pain (Q11-20) | 4.56 | 4.64 | 3.98 | 3.74 |
| General musculoskeletal pain (Q21-30) | 4.66 | 4.72 | 4.10 | 3.86 |

Agreement of the subject matter experts within ±1 point was high across all domains with the majority of scores clustered at the upper end of the 5-point Likert scale, and most responses receiving scores of 4 or 5. *Clarity* had the strongest overall agreement, followed by *Relevance, Completeness*, and *Accuracy (Table 6)*. Most rating differences were limited to a single scale point. Krippendorff’s alpha (ordinal) demonstrated similarly low reliability across domains, with values of −0.04 for Relevance, −0.03 for Clarity, 0.11 for Completeness, and 0.07 for Accuracy. (Table 7).

**Table 6.** Overall Agreement within ± 1 Point.

| *Table 6. Overall Agreement within $\pm 1$ Point* |  |  |  |
| --- | --- | --- | --- |
| **Domain** | **Within $\pm 1$ Agreements** | **Total Pairwise Comparisons** | **% Within $\pm 1$** |
| Relevance | 274 | 300 | 91% |
| Clarity | 284 | 300 | 95% |
| Completeness | 261 | 300 | 87% |
| Accuracy | 226 | 300 | 75% |

**Table 7.** Krippendorff’s Alpha (ordinal) assess inter-rater reliability between five expert reviewers.

| **Domain** | **Krippendorff's Alpha (ordinal)** | **Interpretation** |
| --- | --- | --- |
| Relevance | -0.04 | Systematic disagreement; worse than chance |
| Clarity | -0.03 | Systematic disagreement; worse than chance |
| Completeness | 0.11 | Very low agreement; minimal reliability beyond chance |
| Accuracy | 0.07 | Very low agreement; minimal reliability beyond chance |

## Discussion

Subject matter experts generally rated responses from ChatGPT as accurate and in line with current US chiropractic guideline recommendations. Importantly, the responses included important patient safety aspects of current CPGs such as red-flag screening, imaging recommendations, conservative management strategies, and referral considerations. The responses also frequently volunteered follow-up questions to the original prompt to guide the theoretical patient in conversation with the treating chiropractor and recommended management strategies for relevant conditions.

Despite the alignment with CPGs, readability analysis demonstrated that ChatGPT responses were above recommended patient education reading levels.^30^ Measures of readability indicated that ChatGPT responses correlated to high school to college level reading grades, which exceed cited recommendations that patient-education material should be written at or below a sixth to eight grade level.^30^ This suggest that, though ChatGPT responses aligned with CPGs, they may not be easily accessible for many patients.

Rater agreement was high and within ±1 point across the four domains while chance-corrected reliability metrics demonstrated limited inter-rater reliability. Most disagreements were small. The concentration of score ratings towards the top of the scale (4 and 5) may partially explain this discrepancies of reliability. For kappa-based measures, concentrated marginal distributions can increase estimated chance agreement and yield low coefficient despite high observed agreement, a phenomenon commonly described as the kappa paradox.^31^

Agreements were high across the domains of relevance and clarity, which suggest that the responses were generally seen as appropriate and responsible. Lack of agreement was seen more in the domains of completeness and accuracy, which likely reflected how raters interpreted depth and clinical detail.

The findings concur with prior literature that illustrates how LLMs can achieve moderate to high guideline concordance but may exhibit variability depending on prompt framing. The current study extends this work into the chiropractic field at large, including but not limited to spinal manipulation safety, imaging decision-making, and chronic pain management.

Future research should evaluate larger and more diverse sets of chiropractic questions, compare multiple LLM (Google Gemini, Claude, etc.), and investigate performance in case-based clinical scenarios requiring diagnostic reasoning and treatment planning. Studies that also investigate approaches to improve readability of response for patients are also to be considered. Future AI assessment studies may additionally explore how standardized, structured evaluation frameworks and calibrated scoring tools may improve reproducibility and reliability of expert evaluations of AI responses.

## Limitations

Several limitations should be considered when interpreting these findings. First, ChatGPT responses were generated using a single model version (GPT-5.2) at one point in time. Because large language models are frequently updated, outputs may differ across versions, limiting generalizability to future iterations. Additionally, only ChatGPT was used for this study, and other LLM’s may present different responses Second, one response was generated per question, and all five subject matter experts evaluated that same response. Consequently, consistency across repeated outputs was not assessed, limiting generalizability to other responses generated using the same prompts.

Third, responses were generated in isolated sessions with the conversation being cleared between each response. In real-world patient use, question phrasing or prompting, follow-up dialogue, and contextual refinement may meaningfully influence response quality, depth, and specificity.

As the responses were reviewed by clinicians, and not patients, we did not assess whether patients found the output understandable, appropriate, or useful.

Fourth, ChatGPT was not provided with any prompting beyond the study questions. The LLM model was not instructed to respond from the perspective of a chiropractor or to reference current chiropractic clinical practice guidelines. The exclusion of a prompt may have affected the content of responses in this study.

Fifth, although structured Likert-based rubric was used, scoring across domains required subjective interpretation, raters did not complete an initial calibration exercise using human-written responses designed to align or contradict with the CPGs. We were unable to assess the extent to which raters disagreed on domain ratings due to different interpretations of CPGs, rating instructions, or domain definitions. Lastly, the study evaluated concordance with U.S.-based chiropractic clinical practice guidelines only, and findings may not generalize to other regulatory or international practice environments.

## Conclusions

This study assessed ChatGPT’s responses to chiropractic-related questions posed from a patient perspective. Each response was compared against current CPG recommendations for chiropractic management of neck pain, low back pain, and chronic musculoskeletal pain. These findings suggest that ChatGPT responses frequently incorporated guideline-relevant elements, though the responses were lengthy and scored at reading levels that are more advanced than recommended for patient communications. The assessed domains were rated similarly by subject matter experts, although minor scoring differences and ceiling effects attenuated chance-corrected reliability estimates.

## Data Availability

Access to the data will be made upon reasonable request from the corresponding author.

## Supplementary Files

Supplementary File A - *S*upplementary File A. Queries, ChatGPT Responses, and Guideline Recommendations^16–18^

Supplementary File B – Tables consistent with guideline recommendations that were used for comparison.

## Author Contributions

**Conceptualization:** Alex Pham, Clinton Daniels

**Data Curation:** Alex Pham

**Formal Analysis:** Alex Pham, Alexander Peterson, Clinton Daniels

**Funding Acquisition:** Not applicable

**Investigation:** Alex Pham, Jessica Tom, Aubrianna Jones, Clinton Daniels

**Methodology:** Alex Pham, Jessica Tom, Aubrianna Jones, Clinton Daniels

**Project Administration:** Alex Pham

**Resources:** Alex Pham

**Software:** Alex Pham

**Supervision:** Clinton Daniels

**Visualization:** Alex Pham, Clinton Daniels

**Writing – Original Draft Preparation:** Alex Pham

**Writing – Review and Editing:** Alexander Peterson, Jessica Tom, Aubrianna Jones, Clinton Daniels

## Acknowledgements

We would like to acknowledge the expert panel that assessed the ChatGPT statements: Tyler Barton, Kevin Meyer, Morgan Price, Nikhil Sharma, Christopher Roecker. We would to acknowledge Anna Korpak, PhD and Aaron Baraff, PhD of the Seattle Epidemiologic Research and Information Center (ERIC) for statistical consulting.

